# Autonomous Magnetic Resonance Imaging

**DOI:** 10.1101/2020.03.30.20047191

**Authors:** Keerthi Sravan Ravi, Sairam Geethanath

**Author notes:** **Corresponding author:** Sairam Geethanath, 3227 Broadway, New York, NY, 10027, USA.

## Abstract

Access to Magnetic Resonance Imaging (MRI) across developing countries from being prohibitive to scarcely available. For example, eleven countries in Africa have no scanners. One critical limitation is the absence of skilled manpower required for MRI usage. Some of these challenges can be mitigated using autonomous MRI (AMRI) operation. In this work, we demonstrate AMRI to simplify MRI workflow by separating the required intelligence and user interaction from the acquisition hardware. AMRI consists of three components: user node, cloud and scanner. The user node voice interacts with the user and presents the image reconstructions at the end of the AMRI exam. The cloud generates pulse sequences and performs image reconstructions while the scanner acquires the raw data. An AMRI exam is a custom brain screen protocol comprising of one T_1_-, T_2_- and T_2_*-weighted exams. A neural network is trained to incorporate Intelligent Slice Planning (ISP) at the start of the AMRI exam. A Look Up Table was designed to perform intelligent protocolling by optimising for contrast value while satisfying signal to noise ratio and acquisition time constraints. Data were acquired from four healthy volunteers for three experiments with different acquisition time constraints to demonstrate standard and self-administered AMRI. The source code is available online. AMRI achieved an average SNR of 22.86 ± 0.89 dB across all experiments with similar contrast. Experiment #3 (33.66% shorter table time than experiment #1) yielded a SNR of 21.84 ± 6.36 dB compared to 23.48 ± 7.95 dB for experiment #1. AMRI can potentially enable multiple scenarios to facilitate rapid prototyping and research and streamline radiological workflow. We believe we have demonstrated the first Autonomous MRI of the brain.

## 1. Introduction

The accessibility of MRI as a healthcare service ranges from being prohibitive to scarcely available in developing countries [1]. A World Health Organization metric for evaluating the geographical accessibility of MRI is scanner density, measured as number of scanner units per million people (pmp). Globally, scanner density is imbalanced [4]. India – the second most populous country (1.32 billion) – has a scanner density of less than 1 pmp [2]. Eleven countries in Africa (with populations ranging from 0.735 million to 67.51 million) have no scanners [3].

Lack of educational facilities and/or the high costs involved in imparting technical training has resulted in a lack of skilled manpower required to operate MRI systems in developing countries [4]. While imaging has been shown to increase the utilization of facility-based rural health services and to impact management decisions [4], MRI requires technical expertise to setup the patient, acquire, visualize and interpret data. The availability of such local expertise in certain geographies such as sub-Saharan Africa (SSA) is challenging [3,4]. Most countries in SSA have few or no radiologists, with the majority being deployed in cities and metropolitan areas. Therefore, there is a critical, unmet need to make MRI more accessible globally while controlling the cost factors of an MR exam [1].

Inefficient workflows and usage of MRI result in challenges related to financial and temporal accesses in countries with higher scanner densities than the global average of 5.3 pmp [5]. Overutilization of imaging services is acknowledged in the medical industry, primarily driven by financial incentives of the healthcare system, defensive medicine and patient expectations, to name a few [6]. ‘Protocol creep’ is an example that refers to the practice of performing exams by modifying protocols on a case-by-case basis because a standard catalogue of protocols does not exist. This non-uniformity of protocols increases drastically when multiple departments function under a single system [6]. This results in the subject undergoing exams for longer durations than initially intended and directly translates to larger costs to be borne by the subject and/or other stakeholders. Augmenting human capabilities can tackle these challenges and standardise the practice. Autonomous and time-efficient acquisition, reconstruction and visualisation schemes to maximise MR hardware usage and solutions that reduce reliance on human operation of MR systems could alleviate some of the challenges associated with the requirement/absence of skilled human resource [4,7,8]. Multiple studies have been directed toward intelligent slice planning [9] and protocol optimization to reduce the complexity and burden of performing an MRI exam on the technician. Yang et. al [10] perform automatic slice prescription for brain using a neural network. Soltanian-Zadeh et al. [11] optimize pulse sequence parameters specifically for eigenimage filtering. HeartVista (http://heartvista.ai) and Galen MRI (https://www.galenmri.com/) offer commercial cardiac and one button scan packages respectively. However, to the best of our knowledge, these studies do not provide intelligent protocolling.

Rubin et al. state that managing costs in addition to improving quality and outcomes is critical to maximising the value of an imaging service [12]. This motivation is well captured by the definition of MR value – defined as the ratio of actionable diagnostic information to the costs incurred (including the time involved in acquiring that information) [13]. In this work, we introduce and demonstrate the concept of MR value driven Autonomous MRI (AMRI), building on our preliminary efforts [14,15]. First, the methods presented here transform existing scanners into an Intelligent Physical System (IPS). An IPS is characterised by cognizance, taskability, ethicality, adaptability and its ability to reflect [16] and is capable of performing its task with minimal or no human intervention. Second, a Look Up Table (LUT) is introduced for intelligent protocolling. Also, intelligent slice planning is implemented using a neural network to perform automatic landmarking. Third, the ‘Sitrep’ file standard for communication and recording flow of data and control between AMRI components is introduced. The entire procedure of performing an MR exam is autonomous and thus enables a check on the ‘table time’. An AMRI user does not require any technical knowledge to perform an MRI exam. This is significantly different to a remote exam that would require the presence of a well-trained MR technician or radiologist at a different site, and therefore AMRI mitigates the demand for skilled manpower.

## 2. Methods

All AMRI features were implemented in Python 3.6 [17] and related open-source libraries. The volunteer study was approved by the local institutional review board. AMRI operates in two modes: (i) standard mode – where the ‘user’ was any MR safety aware hospital worker (nurse, for example) administering the scan; (ii) self-administered mode – where the ‘user’ was any MRI safety aware subject intending to undergo the exam.

### 2.1 AMRI Setup

The AMRI setup consists of three components: user node, cloud and scanner. This tri-partite setup allows for a logical partitioning of functionalities. The user node is any smart device that interacts with the user via one or more input modalities. Examples of such input modalities could be interacting via voice (this work), keyboard input, a web-form, integration with health information systems, etc. The cloud is any system with significant compute and storage to primarily perform compute-intensive functions and host the knowledge-base. It chooses acquisition parameters that produce the best contrast whilst meeting SNR and acquisition time criteria (described below) to generate pulse sequences for each scan. It also communicates the user’s commands to the scanner and informs the user about scan progress. The scanner was singly tasked with acquiring raw data from the subject based on the instructions from the cloud. It awaited commands from the cloud and automated the UI operation on the scanner console to initiate MR acquisitions. AMRI morphs the scanner from conventionally being a sophisticated system requiring complex operations (such as slice planning, protocol edits based on SNR, contrast, image visualization, etc.) into only a data sensor. Figure 1 illustrates the flow of operation in a typical AMRI scan.

**Figure 1.**
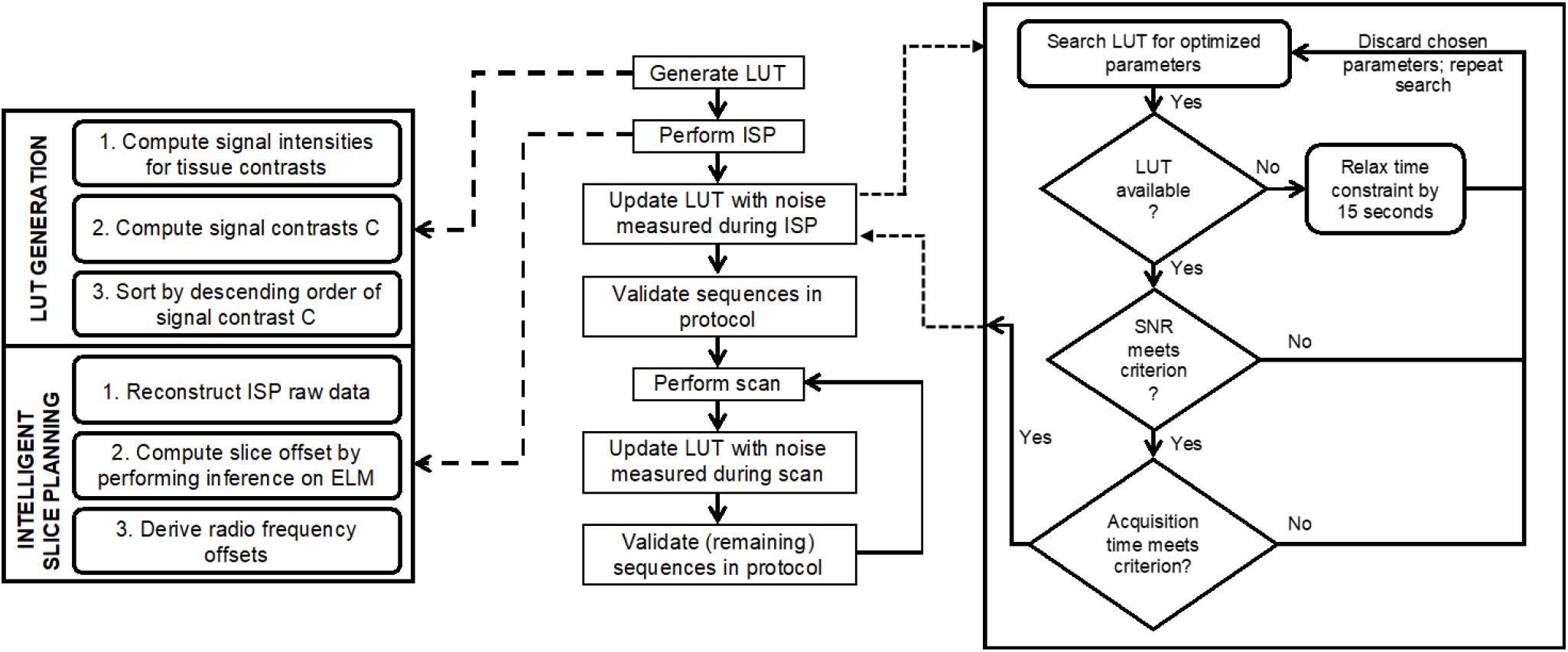
AMRI exam flowchart. A typical AMRI exam includes generating the Look Up Table (LUT), performing Intelligent Slice Planning (ISP) and consulting the LUT to optimise the sequences in the brain screen protocol. The LUT is populated with pulse sequence parameters and signal intensity values derived from standard Gradient Recalled Echo and Spin Echo signal intensity equations. ISP is performed by leveraging a single hidden-layer neural network to predict the slice position of the input axial brain image. Starting with the localiser, the noise measured from each scan is utilised to validate the sequences in the protocol to determine if the SNR and time constraints are satisfied.

### 2.2 Intelligent Slice Planning (ISP)

Slice planning was treated as a multi-class classification problem requiring rapid prototyping and inference. An ELM [18] is a single hidden-layer feedforward neural network that is significantly faster than a traditional feedforward neural network. It demonstrates good generalization performance because it converges on the smallest training error with the smallest norm of weights [19]. The only tunable hyperparameter is the number of nodes and this results in faster prototyping. We chose to implement slice planning using an ELM because of this combination of good generalisation performance, fast learning speed, low memory consumption and easy hyperparameter tuning. The multi-class classification problem was designed as follows (Figure 2): the training dataset consisted of pairs of in-vivo axial brain images and their corresponding slice positions in a brain volume. The trained ELM predicted a slice position (output) of a previously unseen axial brain image (input) during testing. This was used to determine the distance to the chosen landmark as offsets to the radio frequency pulse.

**Figure 2.**
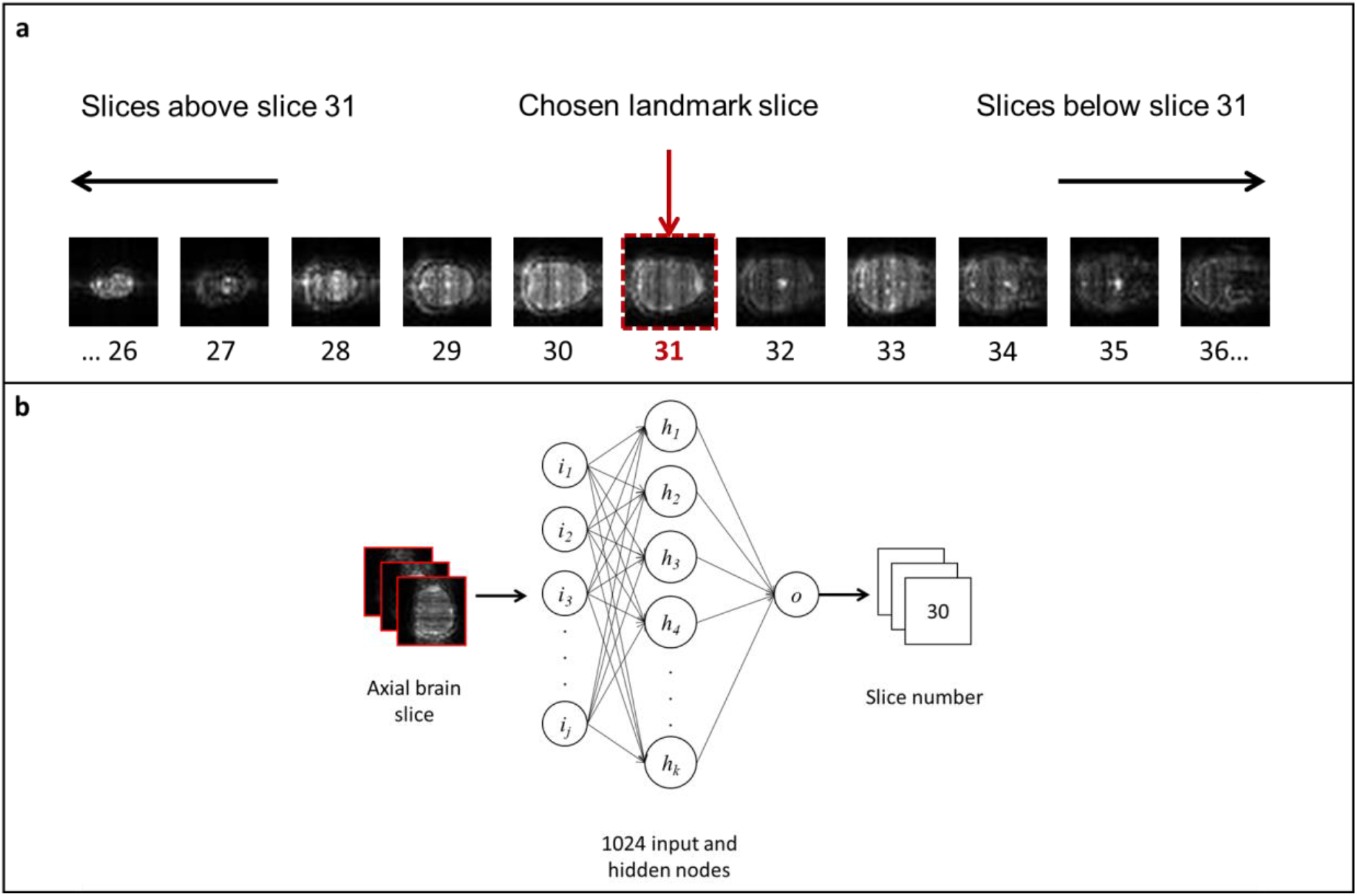
Implementation of Intelligent Slice Planning (ISP). (a) ISP was treated as a multi-class classification problem using an Extreme Learning Machine (ELM). (b) The ELM is trained on pairs of in-vivo axial brain images and their corresponding slice positions in the axial brain volume. The trained ELM is then tasked with predicting the slice position of a previously unseen in-vivo axial brain image, which is then used to determine the radio frequency offsets for slice planning.

In-vivo axial volume data from one human brain was acquired using a standard Gradient Recalled Echo (GRE) sequence to generate the training dataset. The same pulse sequence was also used as the custom localiser at the beginning of every AMRI exam. The acquisition parameters were: TE/TR=8 ms/15 ms, flip angle=56.7°, slice thickness=5 mm, FOV=220 mm and matrix size=32×32. Numpy [20] and Scipy [21] libraries were used to augment the dataset to consist of 5490 samples. First, the acquired slices were rotation-augmented from −30° to +30° in steps of 1° utilizing a bilinear interpolator. This dataset was then replicated three times and noise derived from a uniform distribution scaled by three percent was added. Each data sample was reshaped into a row vector and thresholded to the noise computed earlier. Each row vector was zero-padded to ensure each sample in the dataset was consistently 1024 samples long. The data set was split 90%-10% for training and validation. Since the custom localiser acquired 32×32 axial images, we implemented an ELM consisting of 1024 nodes activated by a sigmoid function and minimised categorical cross-entropy loss. The ELM was trained to achieve a validation accuracy of 87.5% in less than sixty seconds on a 2.5 GHz Intel Core i7, AMD Radeon R9 M370X 2GB Apple MacBook Pro (Apple Inc., Cupertino, USA). The slice offset predicted by the ELM was multiplied by the slice-thickness to derive the radio-frequency (RF) offset. This RF offset was utilized to design the pulse sequences for the subsequent scans.

### 2.3 Intelligent protocolling

Look Up Tables (LUTs) were constructed to accomplish intelligent pulse sequence parameter tuning and adhering to acquisition time constraints (T_max_). One LUT each for the T_1_, T_2_ and T_2_* tissue contrasts was generated. They contained exhaustive combinations of ranges of repetition time (TR), echo time (TE) and flip angle (FA) acquisition parameters based on GRE and SE signal equations [22,23]. These ranges for each of these acquisition parameters were chosen such that the resulting images did not deviate significantly from the desired contrasts. It also contained acquisition times (T_acq_), brain matter signal intensities and contrast values analytically computed for each combination of these parameters. Signal intensities of grey, white and CSF matters were computed as per the standard spoiled-GRE signal intensity equation for each combination, and contrast values were computed as the absolute differences in signal intensities between the appropriate brain matters. The differences were: grey matter from white matter for T_1_, white matter from grey matter for T_2_ and grey matter from CSF for T_2_* contrasts. Each LUT was sorted in descending order by contrast value. A noise value was computed from the acquisition after each scan. Four 10×10 corner patches of the 32×32 image reconstruction from the ISP acquisition were averaged and multiplied by 1.25 to obtain the noise threshold value. This was performed for robustness to include any additional noise components during subsequent pulse sequences. This noise value was used to compute SNR values for each combination of parameters and appended to the LUT. The noise value was computed from the ISP acquisition for the first scan in the protocol. A SNR of 10 dB implies that the signal is approximately three times stronger than the interfering noise. However, a SNR threshold of 10 dB was not achievable with the TR, TE and FA acquisition parameters available in the LUT. Hence, we empirically chose a threshold of 9 dB for the SNR criterion.

The standard AMRI exam involved searching the LUTs to derive the best combination of pulse sequence design parameters for each exam that satisfied the acquisition time constraint (T_max_) and the SNR criterion. The first row of each LUT containing the combination of parameters (TE, TR, FA) producing the best tissue contrasts was chosen at the beginning of the search. If the SNR criterion was met in all three cases, their corresponding acquisition times (T_acq_) were summed, and the exam proceeded if the cumulative acquisition time met the time constraint (∑T_acq_ ≤ T_max_). Otherwise, the LUTs for T_2_ and T_2_* contrasts were alternatively traversed to derive combinations of parameters producing the next-best tissue contrasts while also meeting the SNR criterion. Subsequently, the exam proceeded only if the new cumulative acquisition time satisfied the time constraint (∑T_acq_ ≤ T_max_). If the LUT was exhausted while attempting to derive a combination of parameters that met all criteria, AMRI relaxed the acquisition time constraint by 15 second increments (T_max_ + 15). The LUT was then re-populated and the search was repeated after each increment. The SNR values in the LUTs were updated with noise computed from the image reconstructions of the most-recent acquisition after each scan.

### 2.4 Sitrep and other open source file standards

We propose the Sitrep file standard for communication between the user node, cloud and scanner. Borrowed from military parlance, it is short for ‘situation report’ [24]. Sitrep contains identifying information and records the sequence of events during an autonomous MR exam as key-value pairs. The key identifies the event and the value indicates the state of the event. The vendor-agnostic and open source Pulseq [25–27] file standard (.*seq*) was leveraged by the cloud to configure pulse sequences based on the parameters derived from the LUT. The scanner executed these .*seq* files at the time of data acquisition.

### 2.5 Experiments

In our demonstrations, the user node was an Apple MacBook Pro, the cloud was an Apple iMac Pro, and the scanner was a Siemens Prisma 3T (Siemens Medical Solutions, Malvern, PA). The user node and cloud were connected via the Internet, while the cloud and scanner were connected via a local area network. Three experiments were designed and performed on four subjects to demonstrate AMRI’s cognizance, taskability, adaptability, ethicality and the capacity to reflect (requirements to qualify as an IPS [16]). All the experiments involved acquiring three slices. The experiments differed in the imposed acquisition time constraints (expressed in minutes: seconds).

After every experiment, the patient table position was reset but the patient was not repositioned. The first experiment (#1, 22:30) was designed to demonstrate a scenario in which AMRI could utilize acquisition parameters producing the best contrast while meeting the SNR constraint (corresponding to the ‘best’ choice of parameters). Experiment #2 (13:30) was designed to require AMRI to consult the LUT to derive a combination of parameters that met the two criteria. Experiment #3 (11:30) was designed to force a situation wherein the LUT was exhausted in attempting to derive a combination of parameters that met both the SNR criterion and the time constraint. Therefore, AMRI needed to relax the time constraint in increments of 15 seconds until it derived a choice of parameters meeting the criteria. This experiment was designed to demonstrate cognizance: the system is aware of not meeting the prescribed requirements and attempts to derive a set of working parameters by relaxing a certain condition. Experiment #1 was considered to be the internal control, as the sequences implemented were based on PyPulseq [26] and a vendor-specific interpreter module. Table time was defined as the time spent by the subject in the scanner, inclusive of the communication overheads between the user node, cloud and scanner and the time spent registering the subject.

### 2.6 Data analysis

SNR analysis was performed by taking the ratio of mean signal intensity from the brain images to noise averaged from four 2 × 2 voxel corner patches. The T_1_, T_2_ and T_2_* contrast images were referenced to hand-drawn region-of-interest (ROI) masks for white matter, grey matter and CSF respectively. Tissue matter contrast analysis was performed using these ROI masks to compute absolute differences in signal intensities between white and grey matter (T_1_ and T_2_) and CSF and grey matter (T_2_*). The theoretical maximum and minimum were computed as the ratios of maximum contrast to the smallest acquisition time and minimum contrast to largest acquisition time. MR value for each experiment was calculated as the ratio of cumulative theoretical contrast values (across T_1_, T_2_, T_2_*) to the cumulative acquisition time (∑T_acq_).

## 3 Results

Figure 3 shows the image reconstructions of a representative data set across the three experiments. Figure 4(b-d) are state diagrams that show the activation instants of the user node, cloud and scanner across the three experiments. Experiments one, two and three are colour-coded as green, orange and red respectively. The first instance of activation is when the user node requests the cloud for an encryption key once the user begins registering the subject at the user node. The back and forth communication between the three AMRI components are marked (in line with the steps shown in Figure 1), and the experiments end with the user viewing the reconstructed images on the user node. The acquisition times (minutes: seconds) for experiments #1 (control) and #2 totalled to 22:04 and 13:26, and the table times were 28:16 and 19:51 respectively. Experiment #2 was 29.77% faster than the control. In experiment #3, the time constraint was relaxed from 11:30 to 12:00, and the user’s consent was requested to proceed with the modified time constraint. The acquisition time was 11:56 and the table time was 18:45. Comparing table times, experiment #3 was 33.66% faster than the control, and 5.54% faster than experiment #2. Table 1 lists the contrast values, SNR and acquisition time for the three experiments and three contrasts. Experiments #2 and #3 preserve contrasts similar to control (contrast column means in Table 1). Experiment #3 shows the highest standard deviation compared to control, as is expected due to significant reduction in acquisition time. The SNR values achieved for each contrast across experiments have small standard deviations compared to the mean (SNR column means in Table 1). The SNR values achieved across the experiments have larger standard deviations because the T_1_ contrast produced comparatively smaller SNR. Table 1 also presents MR value computed for the three experiments. Experiment #3 achieves the highest MR value as a result of achieving similar SNR and contrast values while being 33.66% and 5.54% faster than the control and experiment #2 respectively (table time). This demonstrates that AMRI was able to intelligently optimise the protocol to minimise the acquisition time without sacrificing SNR. We observe that ISP achieves similar slices in each of the three experiments even though the position of the patient table was reset at the end of each experiment. This shows the slice offsets from the desired slice location computed by the ELM was different for each of the three experiments (−30, −35, −25) due to different subject positioning whereas the resulting slices in each experiment were similar.

**Table 1.** Quantitative analysis of image reconstructions from three experiments across four subjects. Experiment #3 was 33% faster than experiment #1 and produced an SNR of 21.84 ± 6.36 dB versus 23.48 ± 7.95 dB. Experiment #3 achieved the highest MR value of 1.4 × 10-3. In this work, MR value is defined as the ratio of actionable diagnostic information to acquisition time; in this work we derive MR value as the ratio of sum of all achieved contrast values to the corresponding experiment’s acquisition time (T_acq_). Currently, MR value is defined on an arbitrary scale. Experiment 3 required AMRI to relax the acquisition time constraint (T_max_) by 30 seconds, resulting in a new T_max_ of 12:00. Experiments #1, #2 and #3 are marked by their respective colours from Figure 3.

| | Time constraint ($T_{\max}$, minutes: seconds) | Table time (minutes: seconds) | Acquisition time ($\sum T_{\text{acq}}$, minutes: seconds) | SNR (dB) | | | | Contrast (au) | | | | MR Value (a.u.) |
| --- | --- | --- | --- | --- | --- | --- | --- | --- | --- | --- | --- | --- |
| | | | | $T_1$ | $T_2$ | $T_2^*$ | Mean ±SD (dB) | $T_1$ | $T_2$ | $T_2^*$ | Mean ±SD (a.u.) | |
| Experiment 1 | 22:30 | 28:16 | 22:04 | 14.54 | 29.78 | 26.12 | 23.48 ±8.0 | 0.43 | 0.28 | 0.34 | 0.35 ±0.07 | $0.79 \times 10^{-3}$ |
| Experiment 2 | 13:30 | 19:51 | 13:26 | 15.00 | 28.45 | 26.33 | 23.26 ±7.2 | 0.45 | 0.27 | 0.33 | 0.35 ±0.09 | $1.3 \times 10^{-3}$ |
| Experiment 3 | 11:30 | 18:45 | 11:56 | 14.51 | 25.90 | 25.11 | 21.84 ±6.4 | 0.44 | 0.20 | 0.39 | 0.34 ±0.12 | $1.4 \times 10^{-3}$ |
| Mean ± SD | | | | 14.68 ± 0.3 | 28.04 ± 2.0 | 28.85 ± 0.7 | 22.86 ± 0.9 | 0.44 ± 0.01 | 0.25 ± 0.04 | 0.35 ± 0.03 | 0.34 ± 0.01 | $1.16 \times 10^{-3} \pm 2 \times 10^{-4}$ |

**Figure 3.**
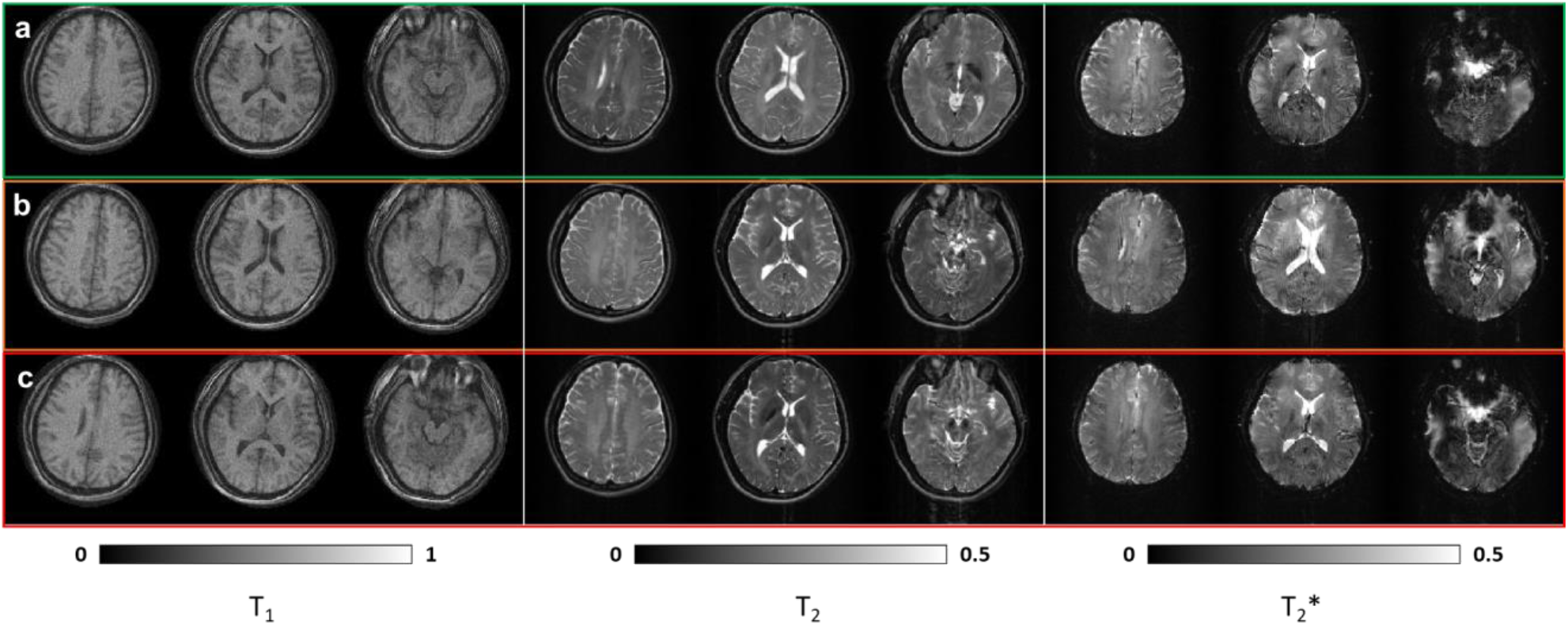
Image reconstruction of the three slices acquired across three experiments from one subject. Experiments #1, #2 and #3 are highlighted by green, orange and red bounding boxes respectively (a)-(c) Reconstructions of data acquired in experiments one, two and three with acquisition time constraints of 22:30, 13:30 and 11:30 respectively. In experiment #3, the time constraint was relaxed to 12:00 to meet SNR criterion. Reconstructions are representative of quality of data acquired from remaining three subjects.

**Figure 4.**
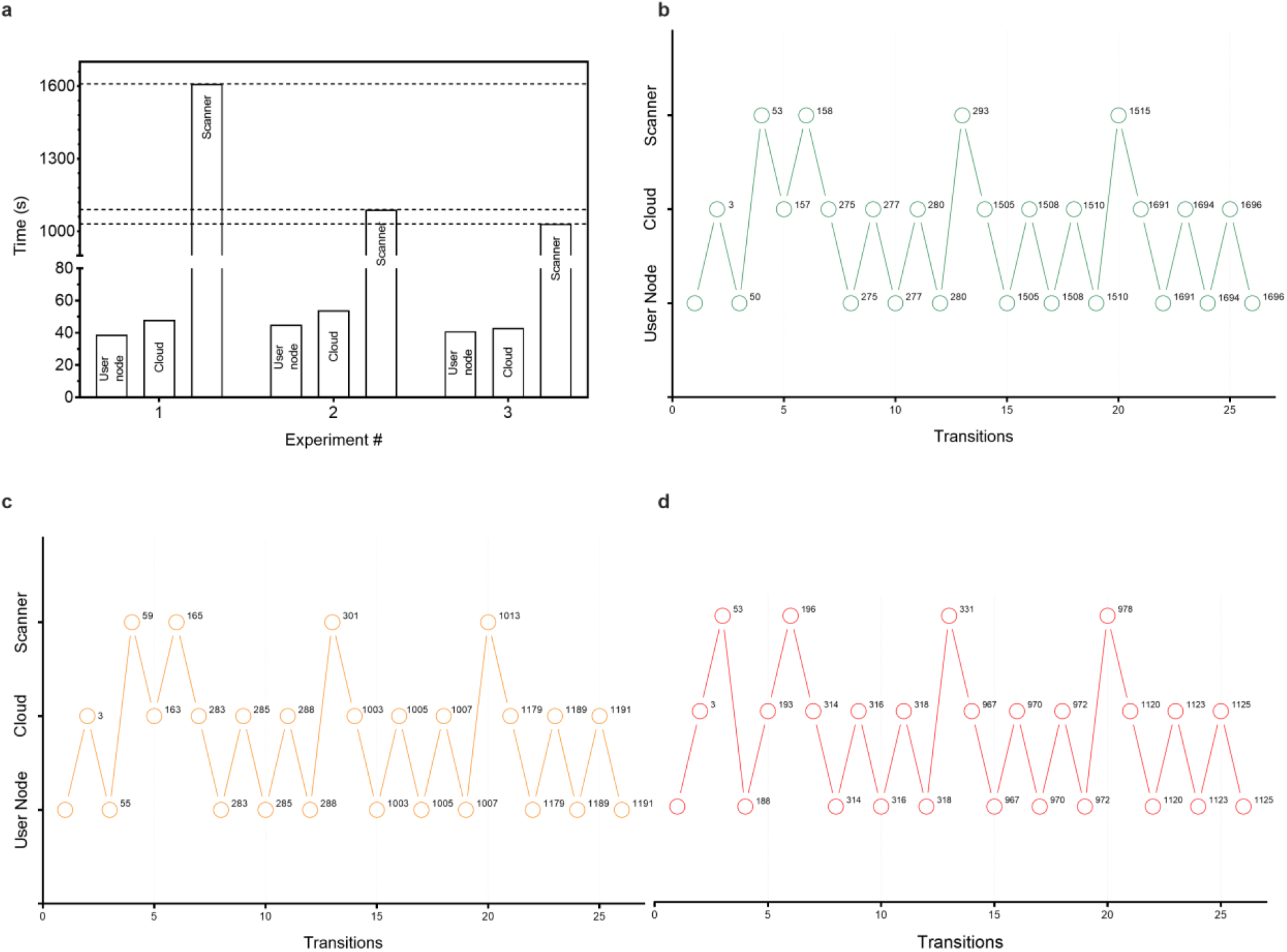
Autonomous MRI component-activity timing diagrams. (a) Time spent in seconds by each component for the three experiments. Time spent at: user node comprises subject registration; cloud includes intelligent pulse sequence design and reconstruction of raw data; scanner comprises acquiring raw data. The dotted lines indicate the acquisition times for each experiment (b)-(d) Activations of the user node, cloud and scanner at different time instants for the three experiments for one subject. Experiments #1, #2 and #3 are colour-coded as green, orange and red respectively. The first activation is the initiation of the registration process at the user node, and the last activation is the cloud uploading the last slice of the last performed scan. The intermediate steps can be referenced from Figure 1.

Figure 4a depicts the cumulative time spent at each component for each of the three experiments. Experiment #3 required the least acquisition time and this is reflected by the shortest scanner bar across the experiments. The user node and cloud bars have similar heights indicating that the table times were primarily affected by the acquisition times. Figure 4(b-d) are state diagrams that show the activation instants of the user node, cloud and scanner across the three experiments. The first instance of activation is when the user node requests the cloud for an encryption key once the user begins registering the subject at the user node. Experiment #3 required obtaining the user’s consent to proceed with modified working parameters; this additional interaction with the user node can be observed as the fourth transition between the scanner and the user node in Figure 4(d).

## 4 Discussion

AMRI’s features are assembled in a novel way that automates the acquisition workflow and transforms a standard MR system into an intelligent scanner. We developed a custom slice planning tool because the vendor-provided feature was not compatible with the PyPulseq pulse sequence programming framework. In this work, we implement MR value as the ratio of theoretical contrast value to the acquisition time. During a standard AMRI exam, the LUTs were sorted in descending order of contrast values, and the acquisition parameters (TE, TR, FA) were chosen so as to satisfy the SNR and time criteria. In this way, AMRI indirectly optimized each MR exam for MR value. Figure 4a depicts the cumulative time spent by each AMRI component during the course of an autonomous MR exam. Figure 4(b-d) are activity timing diagrams for experiments #1-3. Each node indicates a particular step in the AMRI exam, and the number of seconds spent is indicated next to each node. It can be observed that the most amount of time is spent by the scanner as is desirable (Figure 4). All experiments incur an average communication overhead of 32.32% of the total performance time. This can be attributed to the delays incurred in automating the GUI and the length of the file-check intervals when receiving acquired raw data from the scanner. The times indicated in Figure 4(b-d) are inclusive of communication overheads. The current implementation chooses pulse sequence parameters with highest theoretical contrast values subject to SNR and time constraints. However, it is trivial to modify the LUT to optimize for another metric while constraining the remaining parameters. For example, AMRI can be easily modified to optimize SNR subject to contrast and time constraints. Finally, AMRI enables multiple scenarios to provide MRI as an accessible service (Table 2).

### 4.1 AMRI as an Intelligent Physical System (IPS)

In accordance with the definition of an IPS [16], a cognizant MR scanner must be aware of its capabilities and limitations in performing exams and protocols. A taskable MR scanner must be able to interact with the user via one or more input modalities (voice/text/gestures etc.) and interpret possibly high-level and vague instructions. In this work, AMRI was designed to be cognizant of conforming to Signal to Noise Ratio (SNR) and time constraints. As demonstrated in experiment 3, AMRI was aware of not being able to meet the SNR criterion within the imposed acquisition time constraint. Therefore, it relaxed the time constraint in steps of 15 seconds until it could satisfy the SNR and acquisition time criteria. The system subsequently demonstrated taskability by requesting the user’s approval to proceed with the working parameters. Also, AMRI registers subject information via voice interaction with the user and translates that information to influence its subsequent actions related to acquisition. An MR scanner is ethical if it complies with prevailing societal and legal rules and frameworks. As an example, AMRI masks the subject’s name with a unique ID and encrypts the subject’s registration information before uploading it to the cloud for patient privacy. It also leverages a Health Insurance Portability and Accountability Act (HIPAA, https://www.hhs.gov/hipaa/index.html) compliant speech-to-text library to perform the voice interaction. The pulse sequence design tool leveraged in this work implements downstream Specific Absorption Ratio and Peripheral Nerve Stimulation checks to assure patient safety. An adaptable MR scanner must gracefully handle discrepancies encountered. AMRI requests the user to clarify misinterpreted voice commands. It is also able to report to the user in case of demands (with respect to acquisition time) that cannot be met. An IPS MR scanner must also have the ability to reflect and learn from past experiences – own or otherwise. In this work, AMRI tuned pulse sequence parameters for each scan by accounting for the noise measured in the localizer or the previous scan. It also performs Intelligent Slice Planning (ISP) based on the localizer acquisition to image a predetermined location and volume of interest. Therefore, an AMRI scanner belongs to the class of IPS as it possesses all of the above characteristics to function autonomously or with minimal human intervention for long periods of time. Figure 5 maps these characteristics of an IPS to the features of AMRI.

**Figure 5.**
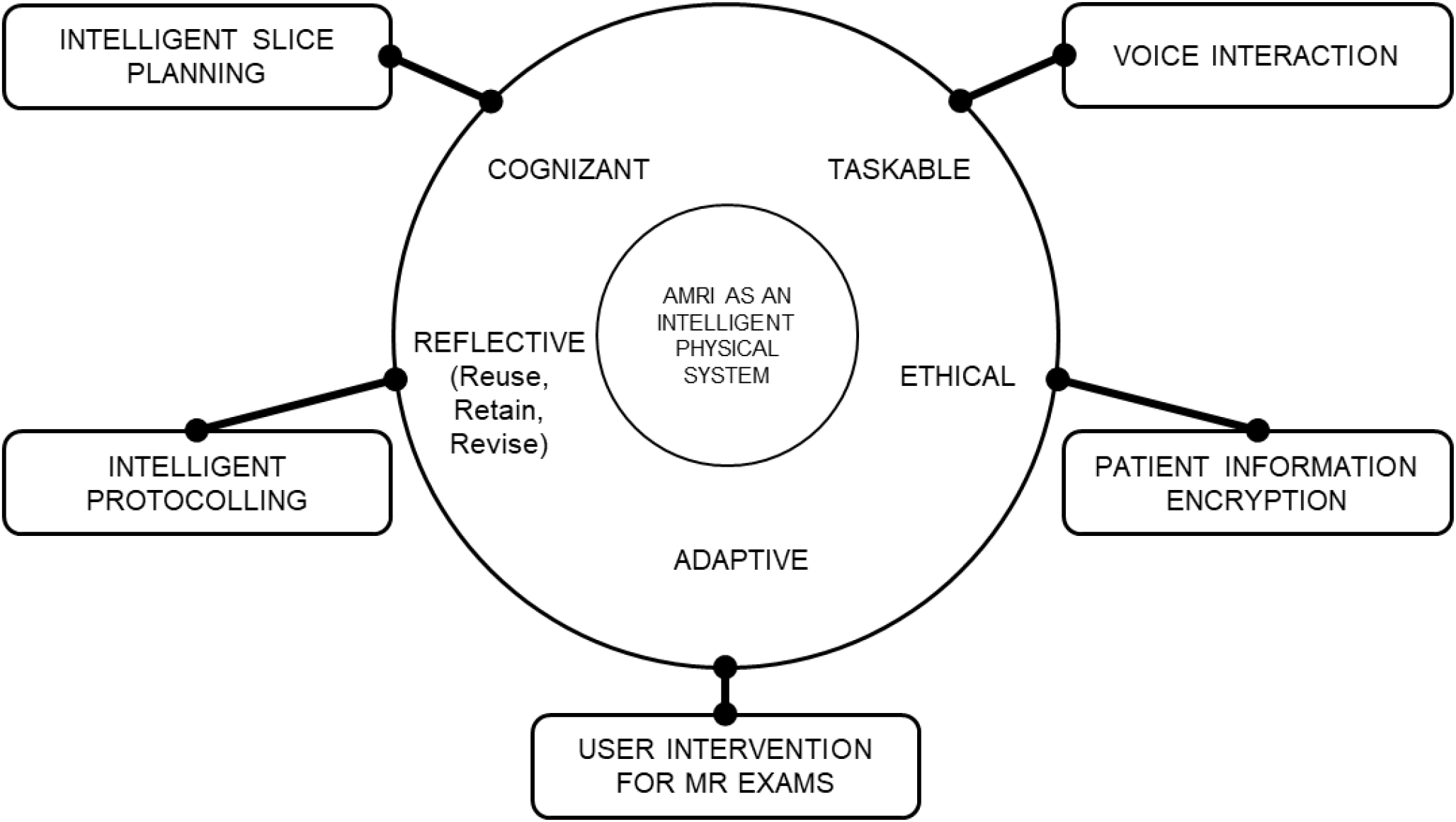
Autonomous MRI (AMRI) as an Intelligent Physical System (IPS). An IPS is characterised as being cognizant, taskable, ethical, adaptive and reflective. Cognizant: AMRI conforms to SNR and time constraints (experiment #3). Taskable: AMRI registers subject information via voice interaction and requests the user’s approval to proceed with the working parameters. Ethical: AMRI masks the subject’s name with a unique ID, encrypts subject information and leverages a Health Insurance Portability and Accountability Act (HIPAA, https://www.hhs.gov/hipaa/index.html) compliant speech-to-text library to perform voice interaction. Adaptable: AMRI requests the user to clarify misinterpreted voice commands. Reflective: AMRI performs slice planning and protocolling intelligently. Therefore, AMRI transforms a standard scanner into an IPS capable of autonomous operation or with minimal human intervention.

An autonomous system performs decision-making processes by optimizing for one or more metrics. This enables the system to take the best decisions to ensure that they are useful and relevant in optimizing for that metric(s). In this work, we tasked the AMRI to optimize for MR value. We implemented and demonstrated a simplified definition of MR value defined as the ratio of contrast achieved to the acquisition time. It is not straightforward to quantify MR value, and hence we simultaneously optimize for SNR and contrast constrained by time spent performing the exam. In a typical AMRI scan, the acquired noise from the ISP acquisition is used to compute the SNR. This enables the LUT to compute contrast values that reflect real-world scenarios. A combination of acquisition parameters that jointly satisfy the SNR, contrast and acquisition time criteria is obtained by searching the LUT.

### 4.2 MRI as an accessible service

Figure 6 illustrates the scenarios made possible by deploying AMRI. This work demonstrates the ‘Remote’ and ‘MR acquisition’ scenarios. The user invoked scans and also viewed reconstructed images on the user node in the ‘Remote’ scenario. The user node and scanner can be in geographically distant locations and communicate via the cloud. Facilitated by Sitrep, the user is updated of the progress of the exam throughout the procedure. The ‘MR acquisition’ scenario enables users without access to MR hardware to upload a ‘.seq’ file generated using PyPulseq to an online form to request acquisition of raw data. The acquired raw data can be reconstructed on the cloud or shared as is with the user.

**Figure 6.**
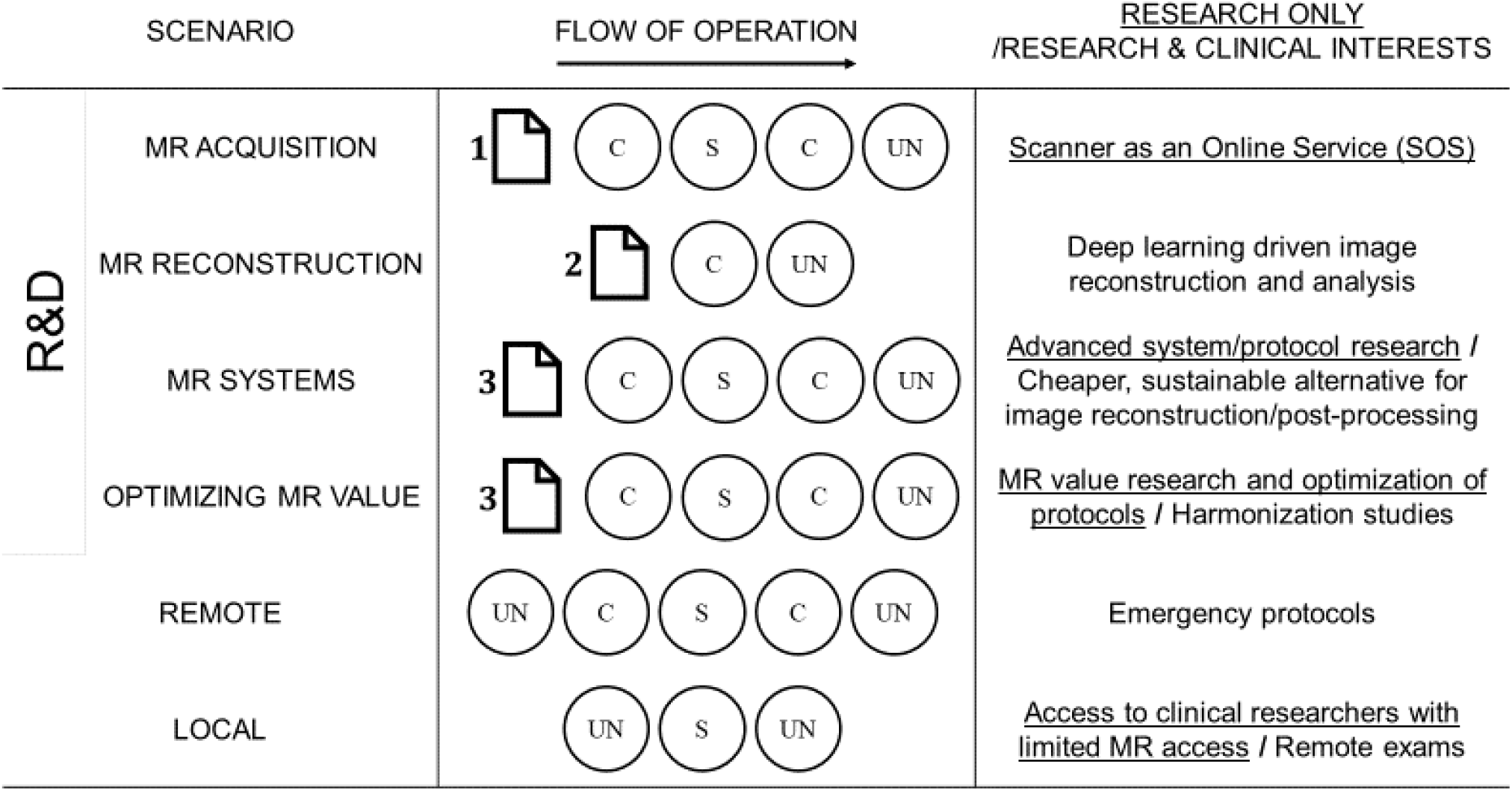
Scenarios enabled by the software introduced in this work. UN, C, and S are abbreviations for user node, cloud and scanner respectively. The scenarios are differentiated by the files and components involved and correspond to different use cases. Files 1, 2 and 3 are pulse sequence exported as a .seq file, raw data in ISMRMRD/DICOM 3.0 format, and Sitrep respectively. This work describes the ‘Remote’ and ‘MR acquisition’ scenarios. The user invoked scans and also viewed reconstructed images on the user node in the ‘Remote’ scenario. The ‘MR acquisition’ scenario enables users without access to MR hardware to upload a .seq file generated using PyPulseq to an online form to request acquisition of raw data. The acquired raw data can be reconstructed on the cloud or shared as is with the user.

The ‘MR systems’ and ‘Optimizing MR value’ scenarios present situations that allow users to rapidly prototype. The method development, scan invocation and image reconstruction can happen on a local cloud (a system with significant compute power and storage installed locally). If such a local cloud is unavailable, a standard system can be used instead. The ‘Local’ scenario is an example of such a situation. Here, the compute-dependent tasks are constrained by the processing power of the available system. To demonstrate some of these scenarios, we adapt AMRI to enable self-administered exams (Section 4.3) and MR scanning as an online service (Section 4.4).

### 4.3 Self-administered exam

With inclusion of inputs from audio-visual (A/V) accessories, the AMRI allowed the user to perform a self-administered brain screen exam [28]. The set up for the self-administered exam utilized a Siemens thirty-two channel DirectConnect head-neck coil and an MR-safe plastic chair. First, the user voice-interacted with AMRI to record registration details. Subsequently, the user landmarked the head-neck coil and then climbed onto the patient table with the aid of the plastic chair as a stepping stool. The user then issued a voice command via MR-safe communication peripherals (OptoAcoustics FOMRI-III+ microphone, OptoAcoustics, Israel) to begin the MR exam. The user was intimated of the progress of the scan via an MR-safe display placed behind the scanner which could be read via a mirror fixed to the head-neck coil. The DirectConnect head coil was setup with the A/V accessories prior to the exam and did not require further manual operation during the exam. The patient table position was moved out to allow the user to exit the scanner at the end of the exam. An illustration of the self-administered MR setup can be found online (http://github.com/imr-framework/amri).

### 4.4 Scanner as an online service

We have also set up an online form (https://forms.gle/J8hi1JaySwqs6HuN9) that enables users to upload a ‘.seq’ file generated using PyPulseq, choose an available phantom, an available receive coil and submit a request to run a scan. We invoke AMRI to obtain the uploaded file, perform the scan and share the raw data with the user at the listed email address. We leverage Google’s Drive online file storage service to receive the uploaded ‘.seq’ files and host the reconstructed images

### 4.5 Limitations

This work was not quantitatively benchmarked against T_1_/T_2_ weighted images from the vendor implementations because PyPulseq’s .*seq* files are vendor-agnostic and we hence chose experiment #1 as our control. Communication overheads and buffer times accounted for 32.32% of the total exam time; subsequent iterations will focus on latency reductions.

#### Intelligent Slice Planning (ISP)

The ELM training dataset is limited in size and diversity, and data augmentation involved only in-plane rotation. The training dataset also did not include oblique slices, and head tilts were controlled for during the experiments. The current implementation does not handle pathological brains. The custom localiser pulse sequence that was implemented has a tight FOV – we suspect it might not generalise well to subjects from diverse backgrounds. Also, a reference coil noise scan was not incorporated. However, such a noise scan does not capture sequence-specific and subject-specific SNR (unless performed for each sequence), which is important to the LUT’s implementation.

#### Look Up Table (LUT)

The LUT does not incorporate a comprehensive definition of MR value. AMRI accomplishes MR value optimization indirectly by attempting to achieve the best contrast within the imposed acquisition time constraint. However, it is not straight forward to optimize for MR value since a globally accepted absolute scale has not yet been defined [7].

Future work involves exploring integration of the vendor-provided slice planning tool to work with the PyPulseq custom pulse sequences. If future iterations continue leveraging the custom slice planning tool, expanding the ELM training dataset to include subjects from varied backgrounds would be beneficial. Professional radiologic assessment of the accuracy of the slice planning would also be beneficial. Reducing the custom localiser’s acquisition time and increasing FOV will make it robust.

## 5 Conclusion

To the best of our knowledge, this is the first demonstration of an MR value driven autonomous scanner. Firstly, this work demonstrates MR value optimized autonomous MRI scanning of the human brain. AMRI qualifies as an IPS by demonstrating the six defining characteristics. Secondly, in-vivo studies of standard and self-administered AMRI exams were performed on four volunteers and similar brain slices were acquired (Figure 3). AMRI optimized to achieve maximum MR value (ratio of image contrast to acquisition time). Thirdly, quantitative analysis of the data acquired from the standard and self-administered AMRI exams was performed (Figure 4). SNR values were consistent within a standard deviation of 3dB and contrast values were consistent within a standard deviation of 0.12 au.

Future work involves protocol optimization and implementing deep-learning solutions to generate a first read radiologist’s report for AMRI; and handling situations where the user intends to perform an emergency exit from the scanner during self-administered MR exams.

## Data Availability

Data will be shared upon request.

## 6 Acknowledgements

The authors wish to thank Jochen Weber for his insights. This study was funded [in part] by the Seed Grant Program for MR Studies of the Zuckerman Mind Brain Behavior Institute at Columbia University (PI: Geethanath).

## Notes

### Competing Interest Statement

The authors have declared no competing interest.

